# Validation of the Hospital Anxiety and Depression Scale in Patients with Decompensated Cirrhosis

**DOI:** 10.1101/2024.07.05.24310010

**Authors:** Chengbo Zeng, John Donlan, Teresa Indriolo, Lucinda Li, Enya Zhu, Joyce C. Zhou, Malia E. Armstrong, Kedie Pintro, Nora Horick, Raymond T. Chung, Areej EI-Jawahri, Maria O. Edelen, Nneka N. Ufere

**Author notes:** Drs. Maria O. Edelen and Nneka N. Ufere contributed equally to this work. **Corresponding Author:** Dr. Nneka N. Ufere Liver Center, Gastrointestinal Unit Department of Medicine Massachusetts General Hospital 55 Fruit Street Boston, MA 02114.

## Abstract

**Background:** The Hospital Anxiety and Depression Scale (HADS) demonstrates strong psychometric properties in many populations of patients with serious illness. However, its psychometric performance among patients with decompensated cirrhosis (DC) has not been examined. We investigated the reliability, validity, and responsiveness of the HADS for patients with DC.

**Methods:** This observational study utilized data from patients with DC at enrollment and week 6 follow-up. Two hundred eighteen outpatients with DC were recruited from a liver transplant center, with 145 completing week 6 assessment. We evaluated psychological distress using HADS and Patient Health Questionnaire 9 (PHQ-9). Patients’ health-related quality of life (HRQOL) was assessed using the Short-Form Liver Disease Quality of Life questionnaire. We examined reliability, floor/ceiling effects, structural validity, and known-groups validity for anxiety (HADS-Anxiety) and depression (HADS-Depression) subscales. We assessed the convergent validity with the PHQ-9. We predicted the change in HRQOL using the change in depression and anxiety. We also evaluated the internal responsiveness to changes in HRQOL for both HADS-Anxiety and HADS-Depression from baseline to week 6.

**Results:** The HADS-Anxiety and HADS-Depression subscales showed strong internal consistency (Cronbach’s alpha>0.8), adequate floor/ceiling effects (<15%), and excellent convergent validity with PHQ-9 (*r*>0.7). Both domains significantly predicted the changes in HRQOL longitudinally. Both HADS-Anxiety (1.8 [95% confidence interval [CI]: 0.5, 3.2]) and HADS-Depression (2.2 [95%CI: 1, 3.4]) showed responsiveness in patients with decreased HRQOL.

**Conclusions:** The HADS is a reliable, valid, responsive tool for assessing anxiety and depression among patients with DC.

## Introduction

Patients with decompensated cirrhosis (DC) experience substantial psychological distress [1,2]. This distress is not only due to the debilitating symptom burden of DC such as ascites, hepatic encephalopathy (HE), and episodes of variceal hemorrhage, but also the psychosocial and financial strain associated with the disease and its treatment [1,3]. Liver transplantation remains the only curative treatment for patients with DC, yet the evaluation process and waiting period are fraught with uncertainties that may further deteriorate patients’ psychological wellbeing. Clinically significant anxiety affects nearly half of patients with cirrhosis, and clinically significant depression affects nearly 1 in 6 [4]. Both depression and anxiety are not only associated with worse health-related quality of life (HRQOL) among patients with DC but also increased mortality [1,5,6]. Therefore, accurate screening of psychological distress among individuals with DC is a critical first step toward reducing their morbidity and mortality.

The Hospital Anxiety and Depression Scale (HADS) is a self-administered questionnaire designed to identify psychological distress in clinical settings [7]. Research has demonstrated that the HADS has strong psychometric properties, making it a valid instrument for evaluating anxiety and depression in a wide variety of patient populations [8–10]. However, research into the psychometric properties of HADS among patients with DC is still scarce. Knowledge of the psychometric performance of HADS would facilitate its application in evaluating anxiety and depression and advancing clinical care for patients with DC. In particular, evidence regarding the validity and responsiveness of HADS in conjunction with HRQOL supports the development of pharmacologic and non-pharmacologic interventions aimed at improving patient outcomes.

The overall goal of this study is to evaluate the psychometric properties of the HADS for patients with DC both cross-sectionally and longitudinally. We hypothesized that the HADS-Anxiety and HADS-Depression subscales would (1) exhibit high reliability, minimal floor and ceiling effects, and robust construct validity; (2) demonstrate good convergent validity with the Patient Health Questionnaire (PHQ-9) and exhibit strong predictive validity in relation to changes in HRQOL; and (3) be responsive to longitudinal changes in HRQOL.

## Methods

### Study Design and Population

This is a secondary data analysis using data of adult patients with DC recruited from the outpatient hepatology clinic at Massachusetts General Hospital between August 2018 and September 2022 [11]. The purpose of the original study is to assess HRQOL, symptom burden, illness and prognostic understanding in a longitudinal cohort of patients with DC. Our eligibility criteria included adult (age ≥ 18 years old) patients who (1) had been diagnosed with cirrhosis complicated by ascites, HE, and/or esophageal variceal bleed (EVB) and (2) were proficient in reading English. We excluded patients with (1) a history of prior liver transplantation; (2) uncontrolled HE, psychiatric disorder, dementia, or other cognitive impairment precluding ability to provide informed consent; (3) hepatocellular carcinoma (HCC) beyond Milan criteria; (4) a current history of extrahepatic malignancy, excluding non-melanoma skin cancer; and (5) those currently receiving palliative care or hospice care.

We collected the following demographic and clinical characteristics from study participants or through a review of the electronic medical record: age, sex, race, ethnicity, relationship status, educational level, liver disease etiology (alcohol, EtOH; metabolic dysfunction-associated steatotic liver disease, MASLD; viral hepatitis B or C; or other), cirrhosis complications (ascites, HE, EVB, and HCC) and severity (Model for End-stage Liver Disease-Sodium score, MELD-Na score; and Child-Pugh class (A [score 5-6]; B [score 7-9]; C [score 10-15]) that were available within two weeks of completion of baseline survey), cirrhosis-specific comorbidity scoring system comorbidity score (CirCom) [12], transplant listing status at the time of enrollment (actively listed for transplant versus not listed).

All research was conducted in accordance with both the Declarations of Helsinki and the Istanbul Study. Study participants did not receive remuneration for study participation. All study participants provided written or verbal informed consent and the Mass General Brigham Institutional Review Board approved this study.

### Patient-Reported Outcomes Measures

Eligible patients had up to 30 days after providing informed consent to complete baseline study questionnaires either verbally (in person or by telephone), on paper, or electronically. Patients who consented were considered enrolled upon completion of baseline study questionnaires, which included the completion of patient-reported outcome measures (PROMs) as described in more detail below. Enrolled patients were contacted at Week 6 after their study enrollment to complete the same set of three PROMs.

#### Hospital Anxiety and Depression Scale (HADS)

We assessed patients’ self-reported anxiety and depression using the 14-item Hospital Anxiety and Depression Scale (HADS) [7]. The HADS has two 7-item subscales assessing the severity of anxiety (HADS-Anxiety) and depression (HADS-Depression) symptoms using a four-point Likert response scale ranging from zero to three. Five of the 14 items were reverse coded, and we calculated HADS-Anxiety and HADS-Depression subscale scores, which range from 0 (no distress) to 21 (maximum distress). Scores higher than 7 on the HADS-Anxiety and HADS-Depression denote clinically significant anxiety or depression.

#### Patient Health Questionnaire-9 (PHQ-9)

The PHQ-9 is a self-administered 9-item instrument used to evaluate depressive symptoms experienced over the past two weeks on a four-point Likert scale (“not at all,” “several days,” “more than half the days,” “nearly every day”) [13]. The total score ranges from 0 to 27, with a higher score indicating a greater level of depression. PHQ-9 scores of 10 or higher denote moderate-to-severe depression [13]. In this study, the PHQ-9 showed strong internal consistency, with Cronbach’s alphas of 0.85 at baseline and 0.89 at Week 6.

#### Short-Form Liver Disease Quality of Life (SF-LDQOL) questionnaire

Patients’ disease-specific health-related quality of life (HRQOL) was assessed using the Short-Form Liver Disease Quality of Life (SF-LDQOL) questionnaire [14,15]. The SF-LDQOL is a 14 question (36-item) scale measuring 9 domains including symptoms of liver disease, effects of liver disease, concentration/memory, health distress, sleep, loneliness, hopelessness, stigma of liver disease, and sexual functioning/problem [14,15]. We scored each item on a Likert scale, normalized these scores to a 0 to 100 range, and then calculated the average score for each domain such that higher scores indicate better functioning in that domain. To determine the overall HRQOL score, we averaged the mean scores from all nine domains. A higher overall score indicates better HRQOL.

#### Statistical Methods

We described the demographic and clinical characteristics for the study sample using median and interquartile range (IQR) for continuous variables, and frequency and percentage for categorical variables. We also described the distribution for each item in the HADS as well as HADS-Anxiety and HADS-Depression subscale scores for patients at baseline.

#### Reliability

To evaluate the reliability of HADS, we calculated Cronbach’s alpha (value ≥ 0.70 indicates good internal consistency) for the HADS-Anxiety and HADS-Depression separately [16].

#### Floor and Ceiling Effects

We evaluated the floor and ceiling effects by calculating the percentage of patients with scores near the minimum and maximum possible scores in each domain, following a prior study [17]. A domain score ranging from 0 - 1 was considered indicative of a floor effect, while a domain score between 20 and 21 was considered indicative of a ceiling effect. We used a threshold of 15% of patients with subscale scores in these ranges to indicate significant floor or ceiling effects [17].

#### Construct Validity

We performed a confirmatory factor analysis with two correlated latent variables (depression and anxiety) to evaluate the structural validity of the HADS. We evaluated model fit using the comparative fit index (CFI; cutoff value >0.95), Tucker-Lewis index (TLI: cutoff value >0.95), root mean square error of approximation (RMSEA; cutoff value <0.06), and standardized root mean square residual (SRMR; cutoff value < 0.08) [18]. Convergent validity was evaluated by calculating the correlations of HADS-Depression subscale score with PHQ-9 score at baseline and Week 6, as well as correlations of change in HADS-Depression domain and PHQ-9 scores from baseline to week 6 follow-up. Because anxiety and depression symptoms often co-exist among patients with DC and may correlate despite being distinct constructs, we also assessed the convergent validity of HADS-Anxiety using PHQ-9 [11]. Correlation coefficients were classified as follows: weak (< 0.4), moderate (0.4 - 0.7), strong (> 0.7) [19]. Since psychological distress is an important component of HRQOL, predictive validity was assessed through a regression analysis examining whether the change in overall SF-LDQOL score was predicted by the changes in HADS-Anxiety and HADS-Depression subscale scores from baseline to Week 6, adjusting for the following potential confounders determined *a priori* given their associations with HRQOL on a review of empirical evidence: age, MELD-Na score at enrollment, presence of ascites or HE, diagnosis of EtOH-related cirrhosis, transplant listing status, presence of HCC, and CirCom comorbidity score [20]. We assessed known-groups validity using analysis of variance to examine whether HADS-Anxiety or HADS-Depression subscale scores at baseline differed based on cirrhosis severity using Child-Pugh classes.

#### Responsiveness

Given this is an observational study, we evaluated the internal responsiveness of HADS-Depression by calculating the mean differences and 95% confidence intervals (CIs) in total scores between baseline and Week 6 for patients who either experienced clinically meaningful improvement or worsening on the PHQ-9 and SF-LDQOL. For the HADS-A, we assessed its responsiveness only using SF-LDQOL as a criterion given that we did not have other PROMs for anxiety. The minimal clinically important difference (MCID) for the PHQ-9 is defined as a change of 5 points and 1.5 points for HADS-Anxiety and HADS-Depression, respectively [21,22]. For SF-LDQOL, we used an MCID cutoff of 8 points based on an ongoing clinical trial on the treatment of refractory ascites using SF-LDQOL as the primary outcome [21]. Responsiveness is demonstrated in these analyses if HADS-Anxiety or HADS-Depression changed significantly corresponding to the clinically meaningful changes in PHQ-9 and/or SF-LDQOL.

Given the minimal amount of missing data for analytic samples at baseline and 6-week assessment, we assumed that the missingness occurred completely at random. Therefore, our analyses were performed using only complete cases. Confirmatory factor analysis was conducted using Mplus version 8.1, while other analyses were conducted using SAS version 9.4.

## Results

### Overview

A total of 218/312 (69.9%) patients consented to participate in the study and completed the baseline survey. A total of 73 patients either died (n=9), underwent liver transplantation (n=11), refused (6), did not receive survey (n=1), or were lost to follow-up (n=46), resulting in an analytical sample of 145 (66.5%) patients at week 6 (Figure 1).

**Figure 1.**
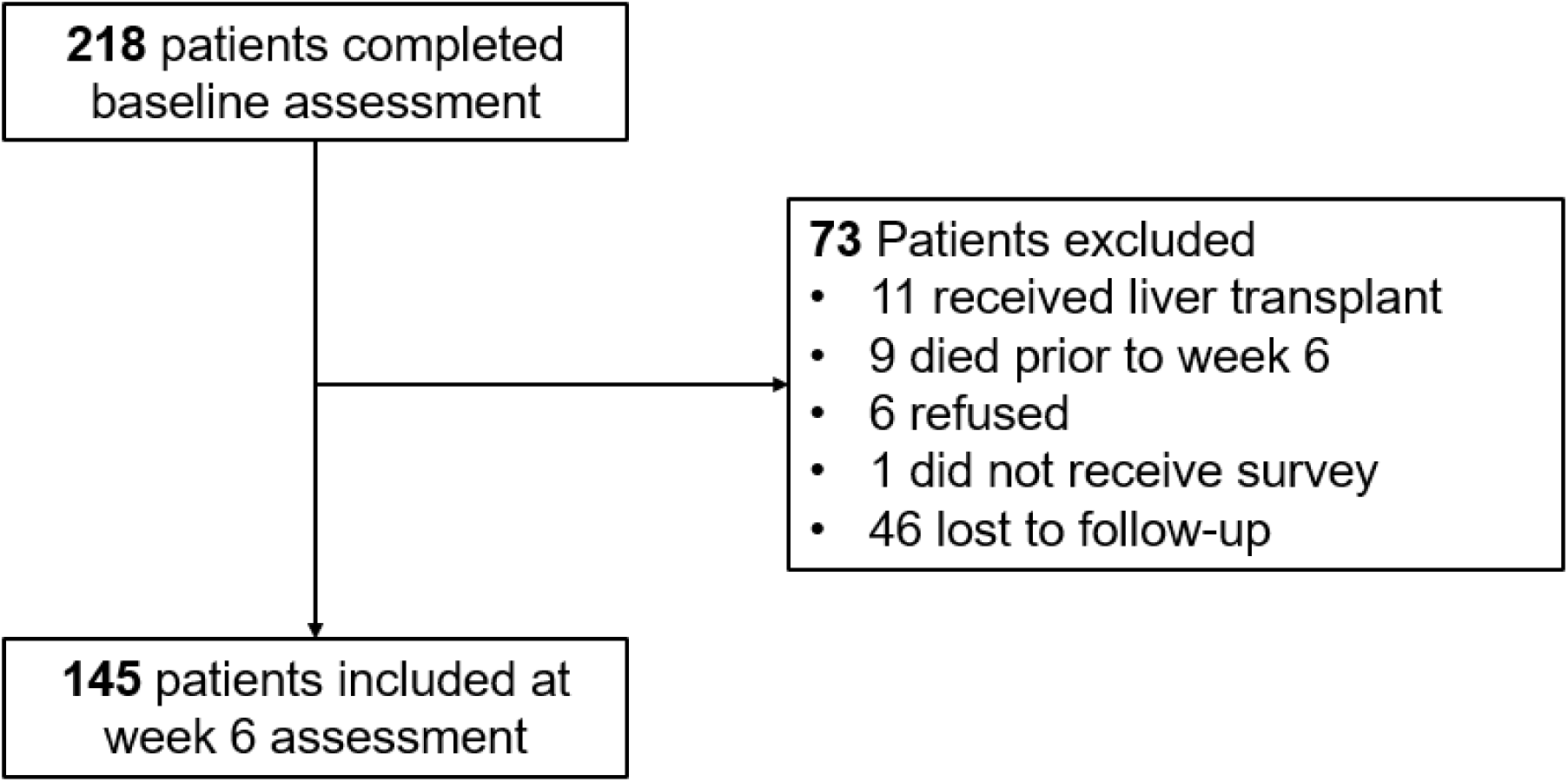
Study inclusion flowchart

In the 218 patients at baseline, the median age was 60 years (IQR: 51 – 65). Most of the patients were White (n=194, 89%), non-Hispanic (n=212, 97%), and either married or living with a partner (n=139, 63.8%). About half of the patients (n=118, 54.1%) were male and actively listed for liver transplantation (n=109, 50%). The median Child-Pugh score was 9 (IQR: 7 – 10), with the distribution of classes as follows: A (n=19, 8.7%), B (n=129, 59.2%), and C (n=70, 32.1%). Table 1 displays the demographic and clinical characteristics for the analytic samples at baseline and at Week 6.

**Table 1.** Demographic and clinical characteristics of the analytic samples.

| **Demographic and clinical characteristics** | **Baseline (N=218)** | **Week 6 (N=145)** |
| --- | --- | --- |
| **Age (median, IQR)** | 60 (51 - 65) | 60 (51 - 66) |
| **Gender** |  |  |
| Male | 118 (54.1) | 81 (55.9) |
| Female | 100 (45.9) | 64 (44.1) |
| **Race** |  |  |
| White | 194 (89.0) | 131 (90.3) |
| African American or Black | 5 (2.3) | 2 (1.4) |
| Other races | 18 (8.3) | 11 (7.6) |
| Missing | 1 (0.4) | 1 (0.7) |
| **Ethnicity** |  |  |
| Hispanic or Latino | 6 (2.8) | 2 (1.4) |
| Not Hispanic or Latino | 212 (97.2) | 143 (98.6) |
| **Relationship status** |  |  |
| Married or living with someone as if married | 139 (63.8) | 97 (66.9) |
| **Educational level** |  |  |
| Bachelor's degree or higher | 86 (39.4) | 66 (45.5) |
| **Liver disease etiology** |  |  |
| EtOH | 127 (58.3) | 82 (56.6) |
| NASH | 50 (22.9) | 34 (23.5) |
| HBV or HCV | 29 (13.3) | 18 (12.4) |
| Others | 12 (5.5) | 22 (15.2) |
| **Decompensation** |  |  |
| Ascites | 200 (91.7) | 132 (91.0) |
| Hepatic encephalopathy | 161 (73.9) | 107 (73.8) |
| Esophageal variceal bleed | 72 (33.0) | 50 (34.5) |
| **CirCom score** | 0 (0 - 1) | 0 (0 - 1) |
| **Child Pugh score (median, IQR)** | 9 (7 - 10) | 9 (7 - 10) |
| A (5-6) | 19 (8.7) | 14 (9.7) |
| B (7-9) | 129 (59.2) | 90 (62.1) |
| C (10-15) | 70 (32.1) | 41 (28.2) |
| **Transplant Status** |  |  |
| Actively listed for transplant | 109 (50.0) | 81 (55.9) |
| Evaluated but not listed | 6 (2.3) | 3 (2.1) |
| No evaluation | 103 (47.7) | 61 (42.0) |
| **HCC at Enrollment** |  |  |
| No | 193 (88.5) | 129 (89.0) |
| Yes | 25 (11.5) | 16 (11.0) |
| **PHQ-9 (median, IQR)** | 8 (4 – 13) | 8 (3 – 13) |
| **HADS-Depression (median, IQR)** | 6 (4 – 9) | 6 (3 – 9) |
| **HADS-Anxiety (median, IQR)** | 7 (4 – 10) | 7 (3 – 11) |
| **SF-LDQOL (median, IQR)** | 57.5 (46.7 – 71.6) | 61.3 (46.8 – 76.5) |

HCC: Hepatocellular carcinoma; CirCom score: cirrhosis-specific comorbidity scoring system comorbidity score; PHQ-9: Patient Health Questionnaire 9; HADS: Hospital Anxiety and Depression Scale; SF-LDQOL: Short-Form Liver Disease Quality of Life

### Item description

The response rates on the 14 items in the HADS were high, ranging from 97% (213/218) to 100% (218/218), as shown in Table 2. The items that received the most responses of 2 or 3 include “I feel as if I am slowed down” (60%), “worrying thoughts go through my mind” (44%), and “I get a sort of frightened feeling as if something awful is about to happen” (36%). Items with positive wording (e.g., ‘I still enjoy the things I used to enjoy,’ ‘I can laugh and see the funny side of things,’ ‘I look forward with enjoyment to things’) had mean scores ranging from 0.5 to 1.2 while items with negative wording (e.g., ‘I feel tense or ‘wound up’,’ ‘Worrying thoughts go through my mind,’ ‘I get sudden feelings of panic’) had mean scores ranging from 0.6 to 1.8. The mean scores for HADS-Depression and HADS-Anxiety were 6.7 (standard deviation [SD]=4.1) and 7.2 (SD=4.3), respectively. In total, clinically significant depression and anxiety were reported by 38.1% (80/210) and 44.1% (94/213), respectively. Eight patients for HADS-Depression and five patients for HADS-Anxiety were excluded due to missingness in HADS items. There were 27.3% (57/209) of the patients with both clinically significant depression and anxiety.

**Table 2.**
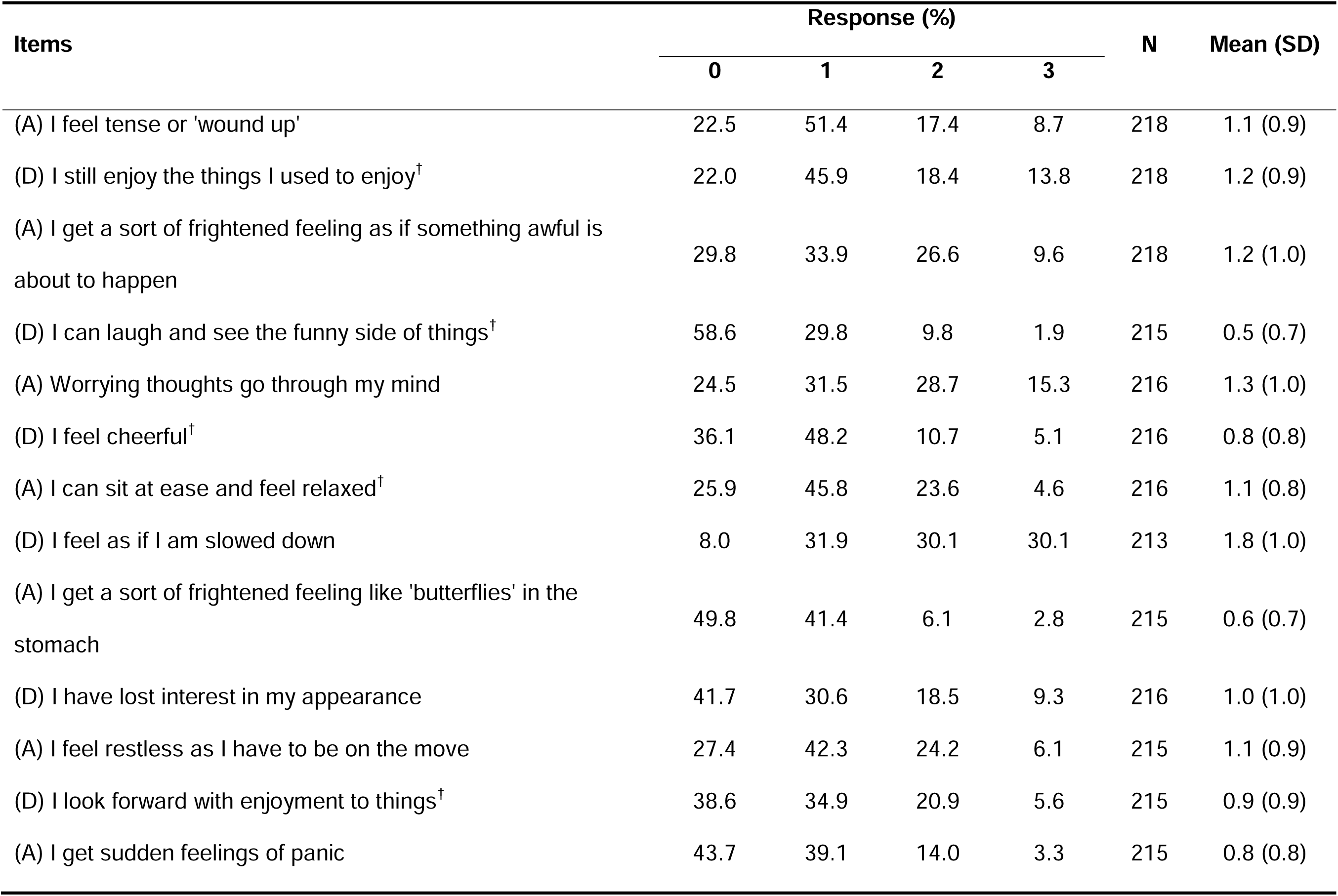
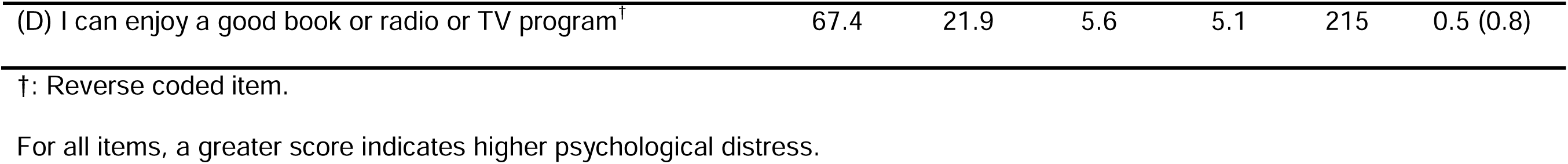
Item description of HADS among 218 patients at baseline.

| Items | Response (%) |  |  |  | N | Mean (SD) |
| --- | --- | --- | --- | --- | --- | --- |
|  | 0 | 1 | 2 | 3 |  |  |
| (A) I feel tense or 'wound up' | 22.5 | 51.4 | 17.4 | 8.7 | 218 | 1.1 (0.9) |
| (D) I still enjoy the things I used to enjoy<sup>†</sup> | 22.0 | 45.9 | 18.4 | 13.8 | 218 | 1.2 (0.9) |
| (A) I get a sort of frightened feeling as if something awful is about to happen | 29.8 | 33.9 | 26.6 | 9.6 | 218 | 1.2 (1.0) |
| (D) I can laugh and see the funny side of things<sup>†</sup> | 58.6 | 29.8 | 9.8 | 1.9 | 215 | 0.5 (0.7) |
| (A) Worrying thoughts go through my mind | 24.5 | 31.5 | 28.7 | 15.3 | 216 | 1.3 (1.0) |
| (D) I feel cheerful<sup>†</sup> | 36.1 | 48.2 | 10.7 | 5.1 | 216 | 0.8 (0.8) |
| (A) I can sit at ease and feel relaxed<sup>†</sup> | 25.9 | 45.8 | 23.6 | 4.6 | 216 | 1.1 (0.8) |
| (D) I feel as if I am slowed down | 8.0 | 31.9 | 30.1 | 30.1 | 213 | 1.8 (1.0) |
| (A) I get a sort of frightened feeling like 'butterflies' in the stomach | 49.8 | 41.4 | 6.1 | 2.8 | 215 | 0.6 (0.7) |
| (D) I have lost interest in my appearance | 41.7 | 30.6 | 18.5 | 9.3 | 216 | 1.0 (1.0) |
| (A) I feel restless as I have to be on the move | 27.4 | 42.3 | 24.2 | 6.1 | 215 | 1.1 (0.9) |
| (D) I look forward with enjoyment to things<sup>†</sup> | 38.6 | 34.9 | 20.9 | 5.6 | 215 | 0.9 (0.9) |
| (A) I get sudden feelings of panic | 43.7 | 39.1 | 14.0 | 3.3 | 215 | 0.8 (0.8) |
| (D) I can enjoy a good book or radio or TV program<sup>†</sup> | 67.4 | 21.9 | 5.6 | 5.1 | 215 | 0.5 (0.8) |

†: Reverse coded item.
For all items, a greater score indicates higher psychological distress.

### Reliability

Both HADS-Depression (Cronbach’s alpha values of 0.80 at baseline and 0.85 at week 6), and HADS-Anxiety (Cronbach’s alpha values of 0.83 at baseline and 0.87 at week 6) demonstrated strong internal consistency.

### Floor and ceiling effects

For HADS-Depression at baseline, the floor effect was 8.1% (n=17) and no ceiling effect was found. For HADS-Anxiety, the floor effect was 9.4% (n=20) and ceiling effect was 0%.

Among patients in Week 6, the floor and ceiling effects for HADS-Depression were 13.8% (n=20) and 0%, respectively. For HADS-Anxiety, the floor and ceiling effects were 13.9% (n=20) and 0.7% (n=1), respectively.

### Structural validity

The confirmatory factor analysis model estimated for HADS fit the data well, as indicated by a CFI of 0.95, a TLI of 0.94, a RMSEA of 0.05, and a SRMR of 0.06. For HADS-Depression, the factor loadings ranged from 0.47 for the item “I feel as if I am slowed down” to 0.74 for both items “I feel cheerful” and “I look forward with enjoyment to things.” For HADS-Anxiety, factor loadings ranged from 0.35 for “I feel restless as if I have to be on the move” to 0.79 for “worrying thoughts go through my mind”. The correlation between two factors was 0.71 (*p*<0.001).

### Convergent validity

At both baseline and Week 6, strong correlations were observed between the PHQ-9 scores and HADS-Depression (baseline: *r* = 0.74; week 6: *r* = 0.82) and HADS-Anxiety (baseline and week 6: *r* = 0.70). Additionally, the change scores for HADS-Depression (*r* = 0.53) and HADS-Anxiety (*r* = 0.45) from baseline to Week 6 demonstrated moderate correlations with the corresponding change scores of the PHQ-9.

### Known-groups validity

There was a clinically significant difference in HADS-Depression scores among patients with varying level of DC severity, both at baseline (Child-Pugh A: 3.8 vs. B: 6.8 and C: 7.5; *p* < 0.001) and Week 6 (Child-Pugh A: 2.1 vs. B: 6.5 and C: 6.7; *p* < 0.001). However, we did not find a clinically significant difference in HADS-Anxiety scores (baseline: Child-Pugh A: 6.6 vs. B: 7.0 and C: 7.7; Week 6: Child-Pugh A: 5.6 vs. B: 6.9 and C: 7.1) (as shown in Figure 2).

**Figure 2.**
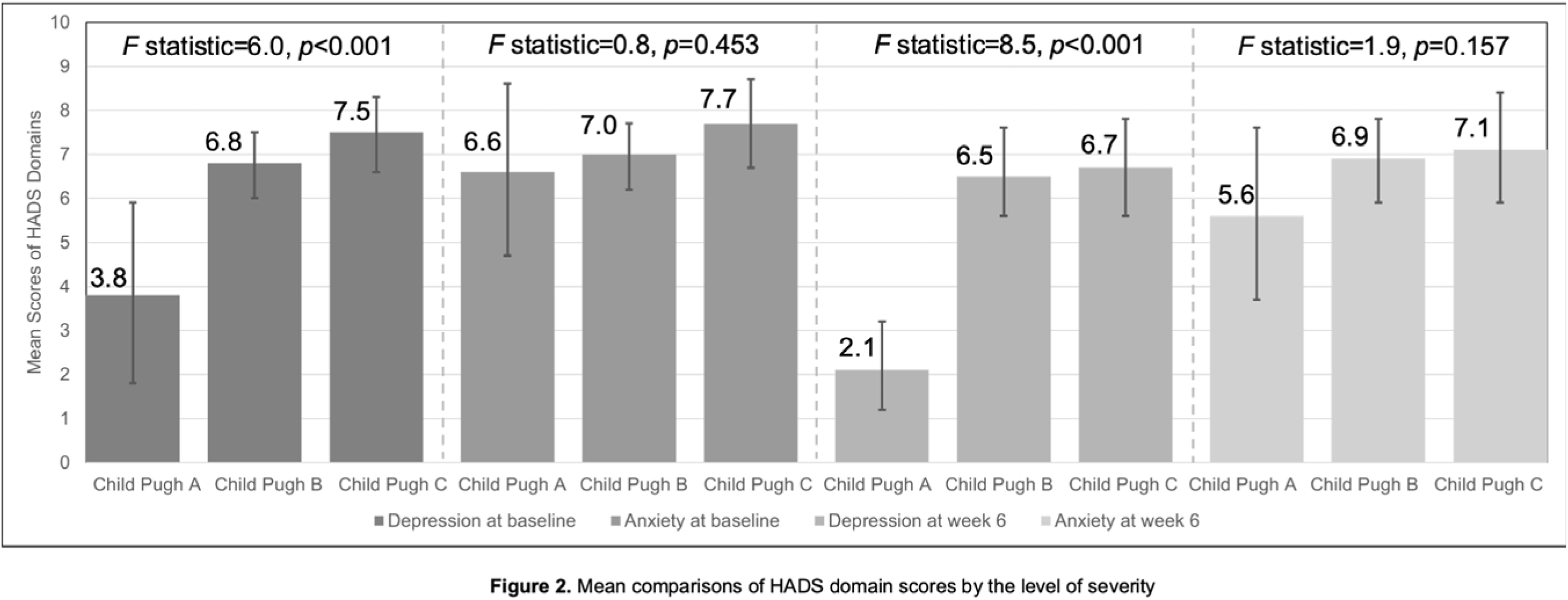
Mean comparisons of HADS domain scores by the level of severity HADS: Hospital Anxiety and Depression Scale. Error bars were the 95% confidence intervals. Eight patients were missing baseline HADS-Depression, and five patients were missing baseline HADS-Anxiety. Two patients were missing HADS-Anxiety in week 6.

### Predictive validity

Among patients who completed the Week 6 assessment, results of regression analysis revealed that the change in HADS-Depression (β=-1.44 [95%CI: -2.06, -0.82], *p*<0.001) was significantly and negatively associated with the change in HRQOL as measured by SF-LDQOL. Similarly, the change in HADS-Anxiety was significantly negatively associated with the change in SF-LDQOL (β=-1.08 [95%CI: -1.60, -0.55], *p*<0.001). Table 3 shows the results of regression analyses.

**Table 3.**
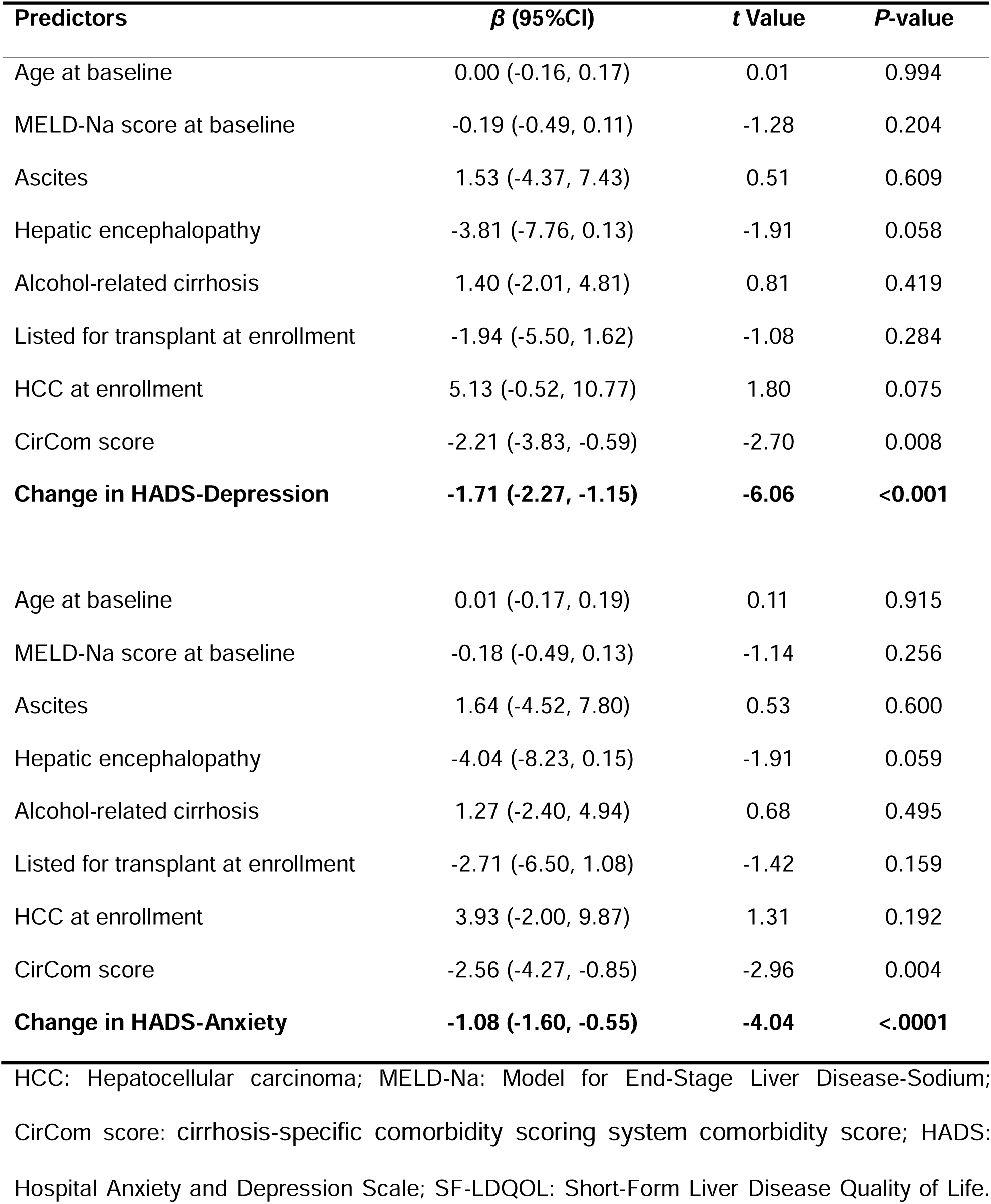
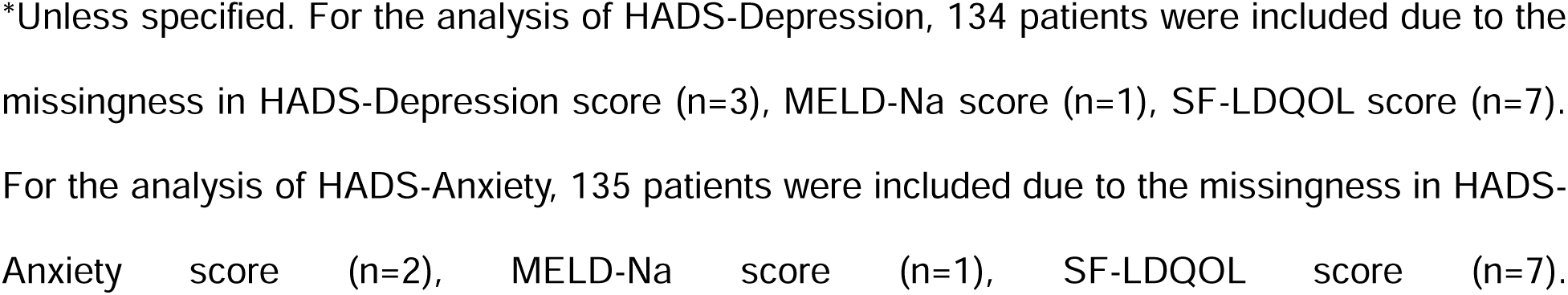
Regression analyses of change in SF-LDQOL predicted by the changes in HADS-Depression and HADS-Anxiety from baseline to 6-week (N=145)*

| Predictors | $\beta$ (95%CI) | t Value | P-value |
| --- | --- | --- | --- |
| Age at baseline | 0.00 (-0.16, 0.17) | 0.01 | 0.994 |
| MELD-Na score at baseline | -0.19 (-0.49, 0.11) | -1.28 | 0.204 |
| Ascites | 1.53 (-4.37, 7.43) | 0.51 | 0.609 |
| Hepatic encephalopathy | -3.81 (-7.76, 0.13) | -1.91 | 0.058 |
| Alcohol-related cirrhosis | 1.40 (-2.01, 4.81) | 0.81 | 0.419 |
| Listed for transplant at enrollment | -1.94 (-5.50, 1.62) | -1.08 | 0.284 |
| HCC at enrollment | 5.13 (-0.52, 10.77) | 1.80 | 0.075 |
| CirCom score | -2.21 (-3.83, -0.59) | -2.70 | 0.008 |
| **Change in HADS-Depression** | **-1.71 (-2.27, -1.15)** | **-6.06** | **<0.001** |
| Age at baseline | 0.01 (-0.17, 0.19) | 0.11 | 0.915 |
| MELD-Na score at baseline | -0.18 (-0.49, 0.13) | -1.14 | 0.256 |
| Ascites | 1.64 (-4.52, 7.80) | 0.53 | 0.600 |
| Hepatic encephalopathy | -4.04 (-8.23, 0.15) | -1.91 | 0.059 |
| Alcohol-related cirrhosis | 1.27 (-2.40, 4.94) | 0.68 | 0.495 |
| Listed for transplant at enrollment | -2.71 (-6.50, 1.08) | -1.42 | 0.159 |
| HCC at enrollment | 3.93 (-2.00, 9.87) | 1.31 | 0.192 |
| CirCom score | -2.56 (-4.27, -0.85) | -2.96 | 0.004 |
| **Change in HADS-Anxiety** | **-1.08 (-1.60, -0.55)** | **-4.04** | **<.0001** |

HCC: Hepatocellular carcinoma; MELD-Na: Model for End-Stage Liver Disease-Sodium; CirCom score: cirrhosis-specific comorbidity scoring system comorbidity score; HADS: Hospital Anxiety and Depression Scale; SF-LDQOL: Short-Form Liver Disease Quality of Life.
\*Unless specified. For the analysis of HADS-Depression, 134 patients were included due to the missingness in HADS-Depression score (n=3), MELD-Na score (n=1), SF-LDQOL score (n=7). For the analysis of HADS-Anxiety, 135 patients were included due to the missingness in HADS-Anxiety score (n=2), MELD-Na score (n=1), SF-LDQOL score (n=7).

### Responsiveness

The HADS-Depression score demonstrated significant changes corresponding to clinically meaningful changes in depression as measured by PHQ-9 from baseline to Week 6, with a mean difference of -4.0 (95%CI: -5.4, -2.6) among patients who showed a decrease of 5 points or more in the PHQ-9 and a mean difference of 2.4 (95% CI: 1.2, 3.5) among those with an increase of 5 points or more in the PHQ-9.

Using HRQOL as a criterion, we found that both HADS-Depression (2.2 [95%CI: 1.0, 3.4]) and HADS-Anxiety (1.8 [95%CI: 0.5, 3.2]) scores increased significantly in patients with clinically meaningful decreases in SF-LDQOL. In patients with clinically meaningful increases in HRQOL, we did not find significant decreases in their HADS-Depression and HADS-Anxiety scores (as shown in Table 4).

**Table 4.** Mean differences in HADS-Depression and HADS-Anxiety between baseline and week 6 by minimal clinically important differences in PHQ-9 and/or SF-LDQOL (N=145)*

|  | Mean difference (95%CI) between baseline and week 6 |  |
| --- | --- | --- |
|  | HADS-Depression | HADS-Anxiety |
| **PHQ-9** |  |  |
| ≤ -5 | **-4.0 (-5.4, -2.6)** | - |
| -4 to 4 | 0.03 (-0.5, 0.5) | - |
| ≥ 5 | **2.4 (1.2, 3.5)** | - |
| **SF-LDQOL** |  |  |
| ≤ -8 | **2.2 (1.0, 3.4)** | **1.8 (0.5, 3.2)** |
| -8 to 8 | 0.0 (-0.6, 0.5) | 0.2 (-0.5, 0.8) |
| ≥ 8 | -0.9 (-2.2, 0.3) | -0.9 (-2.4, 0.6) |

PHQ-9: Patient Health Questionnaire 9; HADS: Hospital Anxiety and Depression Scale; SF-LDQOL: Short-Form Liver Disease Quality of Life. \*Unless specified. For the analysis of PHQ-9 and HADS-Depression, 130 patients were included due to the missingness in change scores of PHQ-9 (n=12) and HADS-Depression (n=3). For the analysis of SF-LDQOL and HADS-Depression, 135 patients were included due to the missingness in change scores of SF-LDQOL (n=7) and HADS-Depression (n=3). For the analysis of SF-LDQOL and HADS-Anxiety, 136 patients were included due to the missingness in change scores of SF-LDQOL (n=7) and HADS-Depression (n=2).

## Discussion

In a cohort of 218 adult patients with DC, we assessed the psychometric validity of the HADS both cross-sectionally and longitudinally. Overall, the HADS exhibited robust structural validity, reflecting the depression and anxiety domains. The HADS-Depression and HADS-Anxiety subscales showed strong internal consistency and adequate floor and ceiling effects. Regarding known-groups validity, we found a clinically significant difference in HADS-Depression when comparing patients with Child-Pugh A disease compared to those with more decompensated disease (Child-Pugh B and C). Both the HADS-Depression and HADS-Anxiety subscales had strong convergent validity with PHQ-9. Notably, both HADS-Depression and HADS-Anxiety had strong predictive validity for longitudinal changes in HRQOL. We further established the responsiveness of HADS-Depression and HADS-Anxiety subscales, which demonstrated both statistically and clinically significant increases in scores among patients with decreased HRQOL longitudinally. Our findings demonstrated strong psychometric properties of HADS in patients with DC and its validity for use in both clinical and research settings.

The robust psychometric performance of the HADS-Depression subscale corroborates its utility as a screening tool among patients with DC. We found that HADS-Depression exhibits strong convergent validity with the PHQ-9 and was responsive to significant changes in the PHQ-9 score over time. Unlike the PHQ-9, which includes items assessing the severity of somatic symptoms that are common among patients with DC such as poor appetite, feelings of tiredness, and sleep disturbance, the HADS-Depression subscale focuses on cognitive and affective symptoms of depression [7,9,13]. In turn, the HADS-Depression subscale may minimize the confounding effects of somatic symptoms. The HADS-Depression for patients at baseline and week 6 displayed an adequate floor effect and a negligible ceiling effect. This is crucial for accurate depression screening in patients with DC, who often experience a spectrum of somatic symptoms that could lead to overestimation of rates of depression on PROMs that assess these symptoms. Therefore, the HADS-Depression subscale, when used alongside other common screening instruments, may more accurately identify patients with DC experiencing depression.

Our results also confirmed the strong psychometric performance of the HADS-Anxiety subscale and its potential as a screening tool for anxiety among patients with DC. Our study found that both the HADS-Anxiety and HADS-Depression domains performed very similarly and demonstrated comparable reliability, structural validity, and convergent validity which aligns with empirical evidence [9]. We further demonstrated that in patients with clinically meaningful decreases in HRQOL, HADS-Anxiety scores increased significantly, establishing its responsiveness among patients with DC. While our results did not demonstrate clinically significant known-group validity for the HADS-Anxiety subscale, we did see appropriate directionality with increasing levels of anxiety among patients with increasing disease severity as assessed by Child-Pugh class.

Our findings also demonstrated the strong relationship between anxiety and depression with HRQOL among patients with DC. Our results indicated that changes in HADS-Anxiety and HADS-Depression scores significantly and independently predicted changes in HRQOL as measured by SF-LDQOL after controlling for other covariates known to affect HRQOL among patients with DC. Psychological health is an important component of HRQOL among patients with DC [14]. However, to date there have been no tested treatment for depression or anxiety among patients with DC [23]. There is a critical need for pharmacologic and non-pharmacologic interventions to treat anxiety and depression in patients with DC to improve their HRQOL.

This study has several limitations. First, we recruited our patients from a single liver transplant center who needed to be proficient in reading English and most of them were non-Hispanic White, limiting the generalizability of our findings. Second, only patients who did not experience liver transplantation and were alive were invited to participate in the Week 6 assessment. Caution is needed when interpreting the results from the Week 6 assessment given the potentially different demographic and clinical characteristics. Third, we could not evaluate the test-retest reliability of the HADS subscale scores in our sample given the long period between baseline and Week 6 follow-up. Future studies are encouraged to assess the test-retest reliability over a short period. Finally, we did not have a second measure of anxiety to evaluate the convergent validity of HADS-Anxiety, but our results found that HADS-Anxiety behaved similar to HADS-Depression when evaluating their convergent validity with PHQ-9.

### Conclusion

The HADS-Depression and HADS-Anxiety subscales within HADS are reliable, valid, responsive tools for assessing psychological distress among patients with DC. Future directions involve the use of HADS in both clinical research and routine screening in cirrhosis care.

## Author’s contributions

CZ, RTC, AE, MOE, and NNU were involved in the concept design and conduct of the study. JD, TI, LL, EZ, JCZ, MEA, and NNU helped with patient recruitment and data curation. CZ, KP, NH, MOE, and NNU performed the data analysis. CZ, MOE, and NNU wrote the initial draft and revised the manuscript. All authors were involved in interpreting the data and providing critical input regarding the analysis and manuscript. All authors approved the manuscript.

## Data availability

The data presented in this study are available on request from the corresponding author.

## Ethical approval

The Mass General Brigham Institutional Review Board approved this study.

